# Multi-omics integration prioritizes potential drug targets for multiple sclerosis

**DOI:** 10.1101/2024.09.26.24314450

**Authors:** Yuan Jiang, Qianwen Liu, Pernilla Stridh, Ingrid Kockum, Tomas Olsson, Lars Alfredsson, Lina Marcela Diaz-Gallo, Xia Jiang

## Abstract

**Background and Objectives:** Multiple sclerosis (MS) is a chronic autoimmune disease with limited treatment options. Thus, drug discovery and repurposing are essential to enhance treatment efficacy and safety.

**Methods:** We obtained summary statistics for protein quantitative trait loci (pQTL) of 2,004 plasma proteins and 1,443 brain proteins, a genome-wide association study (GWAS) of MS susceptibility with 14,802 cases and 26,703 controls, and expression quantitative trait loci (eQTL) for 8,000 genes in peripheral blood and 16,704 genes in brain tissue. Our integrative analysis included a proteome-wide association study to identify MS-associated proteins, followed by summary-data-based Mendelian randomization (SMR) to determine causal associations. We used the HEIDI test and Bayesian colocalization analysis to distinguish pleiotropy from linkage. Proteins passing SMR, HEIDI, and colocalization analyses were considered potential drug targets. We further conducted pathway annotations, protein-protein interaction (PPI) network analysis, and examined mRNA levels of these targets.

**Results:** We identified hundreds of MS-associated proteins in plasma and brain, confirming the causal roles of 18 proteins (nine in plasma and nine in brain). Among these, we found 78 annotated pathways and 16 existing non-MS drugs targeting six proteins. We also discovered intricate PPIs among seven potential drug targets and 19 existing MS drug targets, as well as PPIs of four targets across plasma and brain. Combining expression data, we identified two targets adhering to the central dogma of molecular biology.

**Discussion:** We prioritized 18 potential drug targets in plasma and brain, elucidating the underlying pathology and providing evidence for drug discovery and repurposing in MS.

## Introduction

Multiple sclerosis (MS) is an autoimmune inflammatory disease characterized by demyelination and degeneration in the central nervous system, and has caused non-traumatic disabilities in millions of individuals globally. Disease-modifying therapy (DMT) plays an important role in MS management by reducing the occurrence and severity of relapses and postponing disability accumulation.^1^ Nonetheless, current therapeutic options remain limited and lack targeted specificity, particularly for progressive MS.^2^ Moreover, side effects such as an increased risk of infection present serious concerns and can lead to non-adherence with long-term DMT treatment.^3^ Therefore, continuous exploration of drug discovery, development, and repurposing has become increasingly crucial for improving both the efficacy and the safety of MS treatment.

Proteins serve as the main molecular agents of cellular and biological processes and are therefore considered highly effective drug targets. Compared to genes and transcripts, proteins as end products play a more direct and influential role in determining phenotypes. Studies have demonstrated that although the human genome contains approximately 20,000 protein-coding genes,^4^ the number of proteoforms (distinct protein isoforms) could exceed a million.^5^ Furthermore, the difference between mRNA and protein levels can reach up to 20-30 fold.^6^ On the other hand, even in the absence of evidence at the genomic or transcriptomic levels, proteins can still contribute to diseases, such as through post-translational modifications.^7^ Therefore, direct measurement of proteins not only helps elucidate disease pathology, but also provides potential targets for drug discovery and repurposing.^8^ Proteomic techniques have been used in MS research since the 2000s, and have detected a substantial number of MS-associated proteins.^8, 9^ However, a challenge that is common to these studies is the limited sample size – the involvement of often less than 100 individuals for a panel of thousands of proteins usually yields a huge statistical burden and inconsistent results.

Integrating proteomic data with genetic information forms an effective strategy to unveil the association between protein expression levels and MS development. This approach has been proven promising, as investigational drugs with supportive evidence from genetics are almost twice as likely to succeed in phase II trials (73% vs. 43%)^10^ and to achieve market approval.^11^ Transcriptomic data provides an additional layer of insights by revealing the effect of mRNA expressions on MS. Through investigating transcriptome profiles, studies have identified candidate drugs across a wide range of diseases.^12, 13^ Integrating multi-omics data including genomics, transcriptomics, and proteomics with phenotypic information, potential drug targets can be prioritized with uncovered underlying biological mechanisms.

Complex diseases often manifest in specific tissues, despite genetic risk variants being present in all cells. By targeting relevant and functional tissues, trait-associated molecular agents can be more accurately prioritized with the revealed underlying pathological mechanisms. For MS, while plasma proteins reflect circulatory function and are easily accessed in clinical practice, brain proteins directly mirror MS pathophysiology. Therefore, incorporating both plasma and brain proteomes would aid in detecting disease-relevant signals.

Previous studies have identified several promising findings through integrating strategies. In plasma, a study identified the proteins *FCRL3*, *TYMP*, and *AHSG* as potential drug targets for MS treatment.^14^ These results were complemented by other studies that identified additional proteins, including *CD40*, *TNFRSF1A*, *CD58,*^15^ *PLEK*, *CR1,* and *CD59.*^16, 17^ To date, only one study has focused on brain tissue, the primary affected tissue in MS, identifying 34 associated proteins.^18^ This study only determined the association utilized proteome-wide association study (PWAS). However, the causal relationships, and the potential for pseudo-links due to SNPs linkage disequilibrium (LD) remain to be examined in brain tissue. In our study, we conducted an integrative analysis using the hitherto largest summary statistics of expression quantitative trait loci (eQTL), protein quantitative trait loci (pQTL), and genome-wide association study (GWAS) of MS susceptibility. We utilized cutting-edge statistical approaches to detect potential causal proteins from complex associations and to distinguish pleiotropy from linkage. By integrating tissue-specific pQTL and MS GWAS data, we prioritized 18 potential drug targets (nine in plasma and nine in brain). Combining eQTL data, we identified two of these 18 targets adhering to the central dogma of molecular biology (genetic locus → mRNA transcription → protein translation → MS). We further found 78 annotated pathways and 17 existing non-MS drugs targeting six of the 18 identified potential targets. We also detected comprehensive protein-protein interactions (PPIs) between 7 of the 18 identified potential drug targets and 19 existing MS drug targets. Moreover, we identified PPIs among four potential drug targets across plasma and brain **(Figure 1)**.

**Figure 1.**
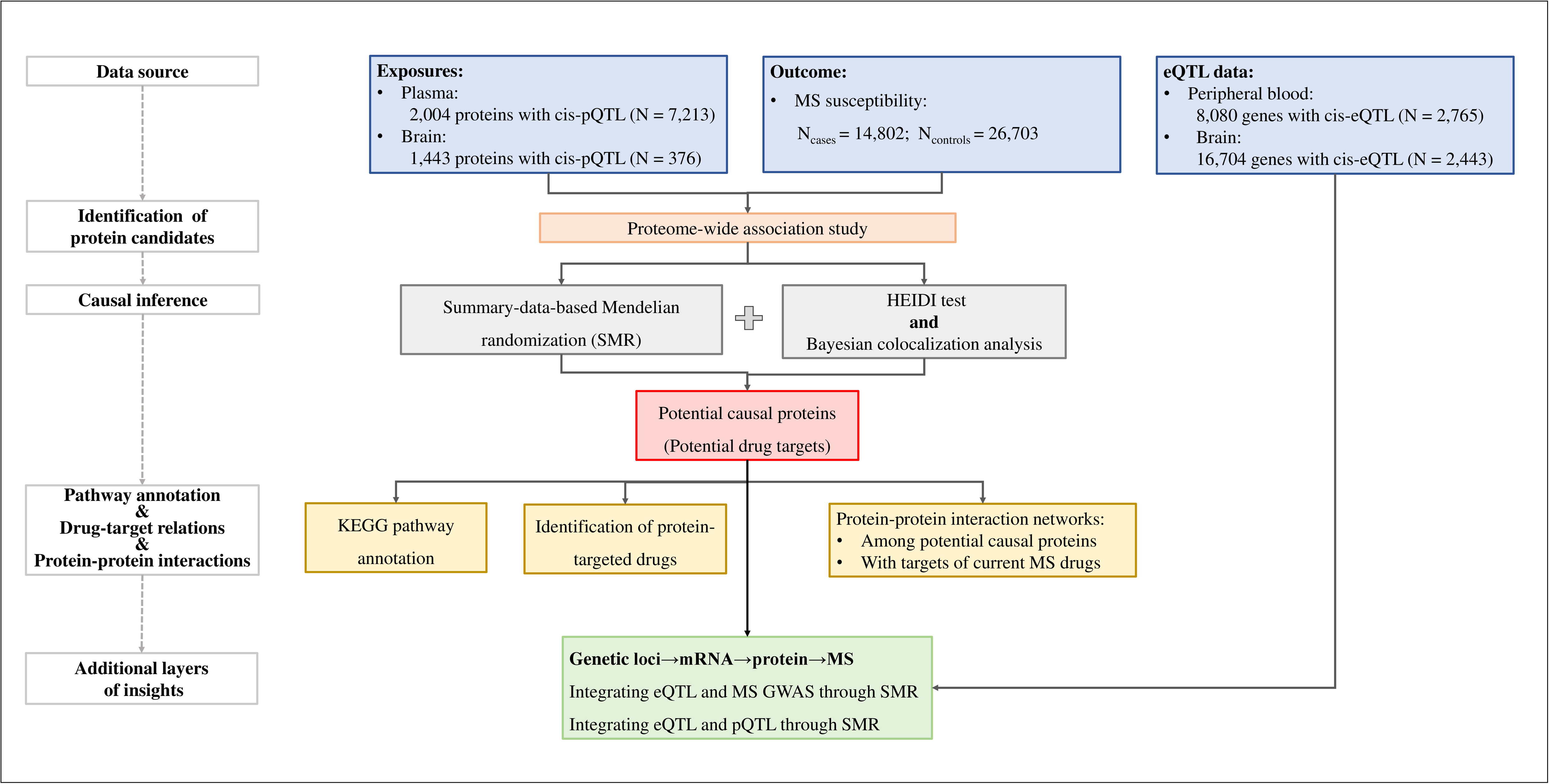
Flowchart of the overall study design. To identify proteins as potential drug targets for multiple sclerosis (MS) treatment, we first conducted a proteome-wide association study (PWAS) to detect proteins associated with MS susceptibility. For candidate proteins identified by PWAS, we conducted a summary-data-based Mendelian randomization (SMR) to test for pleiotropic associations between protein levels and MS. We also performed the HEIDI test and Bayesian colocalization analysis to distinguish pleiotropy from genetic linkage. Proteins that passed all three analyses (SMR analysis + HEIDI test + colocalization analysis) were identified as potential causal proteins. For all detected potential causal proteins, we conducted protein pathway annotation, identified drugs targeting these proteins, and analyzed protein-protein interactions among themselves, as well as interactions with protein targets of existing MS drugs. To gain additional layers of insights, we further investigated mRNA levels of these proteins to assess adherence to the central dogma of molecular biology (genetic locus→mRNA transcription→protein translation→MS). For the identified potential causal proteins, we first analyzed the corresponding mRNA expression levels in relation to MS through SMR analysis (integrating eQTL and MS GWAS). Subsequently, we examined the link between mRNA expression and protein expression through SMR analysis, investigating if a single variant influenced both levels of expression (integrating eQTL and pQTL). By combining the findings from these two analyses, we pinpointed the potential causal proteins following the central dogma. pQTL: protein quantitative trait loci; MS: multiple sclerosis; HEIDI: heterogeneity in dependent instruments test; eQTL: expression quantitative trait loci.

## Materials and methods

### Plasma pQTL summary data

Summary statistics of plasma pQTL were obtained from a large-scale community-based cohort study involving 7,213 participants of European ancestry.^19^ Relative plasma protein concentrations were measured using modified aptamers (SOMAmer). Following quality control for protein-gene mapping, 4,657 SOMAmers with tagged proteins encoded by 4,435 genes were identified. Genotyping was conducted using the Affymetrix 6.0 DNA microarray and subsequently imputed with the TOPMed reference panel. After quality control for imputation quality, Hardy-Weinberg equilibrium, and minor allele frequencies (MAF), 6,181,856 SNPs were identified.

To identify pQTL, a linear regression model was performed, adjusting for age, sex, study site, and ten genetic principal components (PCs). Cis-regions were defined as ±500 KB of the transcription start site. In total, 2,004 significant proteins with cis-pQTLs were identified.

### Brain pQTL summary data

Summary statistics of brain pQTL were obtained from two clinical-pathologic cohort studies using dorsolateral prefrontal cortex of postmortem brain samples donated by 400 participants of European ancestry.^20^ The brain proteome was profiled using liquid chromatography coupled to mass spectrometry analysis. Following quality control for outliers, missing values, and protein loadings, 8,356 brain proteins with corresponding quantitation were detected. Genotyping was conducted either by whole-genome sequencing or by genome-wide genotyping using Illumina OmniQuad Express or Affymetrix GeneChip platforms. Imputation was performed using the 1000 Genomes (1KG) reference panel. After quality control on variants missing rate, Hardy-Weinberg equilibrium, imputation quality, MAF, and degree of kinship, 1,190,321 SNPs on autosomes matching the HapMap LD reference panel were identified.

To determine pQTL, a linear regression model was performed. Cis-regions were defined as a genomic window of 1MB (±500 KB) around genes. In total, 1,443 significant proteins with cis-pQTL were identified.

### GWAS of MS susceptibility

Summary statistics of MS susceptibility were obtained from a recent GWAS including 14,802 MS cases and 26,703 non-MS controls of individuals of European ancestry.^21^ Each of the 15 participating datasets underwent quality checks, followed by imputation using BEAGLE or 1KG reference panel. Subsequently, the association between genetic variants and MS risk was estimated for each dataset through logistic regression, adjusting for the first five PCs. Finally, a fixed-effect meta-analysis was conducted to pool the results.

### Blood eQTL summary data

Summary statistics of peripheral blood eQTL were obtained from the Consortium for the Architecture of Gene Expression, using whole blood of 2,765 individuals of European ancestry.^22^ Gene expression quantification was conducted using Illumina Whole-Genome Expression BeadChips. After quality control on standardizing the expression levels across samples and adjusting for covariates, the mRNA levels for 36,778 transcript expression traits (probes) were investigated. Genotyping was conducted using various platforms and subsequently imputed using the 1KG reference panel. After standard quality control on variants missing rate, Hardy-Weinberg equilibrium, imputation quality, and MAF, 7,763,174 autosomal SNPs were detected.

To identify SNP-probe associations, a linear mixed regression model was performed, adjusting for population structure. Subsequently, Conditional & joint association analysis was conducted to refine the extensive set of identified SNP-probe association results. In total, 11,204 cis-eQTLs for 8,080 genes were identified.

### Brain eQTL summary data

Summary statistics of brain eQTL were accessed from seven cohorts using brain cortex samples donated by 2,443 individuals of European ancestry.^23^ Gene-level transcriptional abundances were quantified using RNA-SeQC. After quality control, retaining individuals with over 10 million reads and an RNA integrity number > 5.5, RNA-seq data from 2,865 brain cortex samples were retained. Genotyping was conducted using various platforms and imputed using the 1KG reference panel and then filtered with standard quality control. In total, 11,631,763 SNPs were detected.

To identify brain eQTL, a linear regression model was performed, adjusting for five genetic PCs. A meta-analysis was then conducted to pool the results across cohorts. In total, 1,962,048 cis-eQTLs for 16,704 genes were identified.

### Statistical analysis

#### PWAS identifying proteins associated with MS, by integrating pQTL and MS GWAS

We first conducted a PWAS to identify candidate proteins associated with MS. FUSION was performed using pre-computed elastic net model-based weights for both plasma and brain protein expressions.^19, 20^ Briefly, the 1KG reference panel was used to reduce the impact of LD on MS GWAS. Subsequently, SNP effect sizes (z-scores) of MS GWAS were imputed using the ImpG-Summary algorithm.^24^ Finally, the association between protein weights and MS GWAS was estimated through a linear regression model. To identify the largest possible number of candidates, a nominal *P*-value < 0.05 was utilized as a significance threshold. Moreover, FDR-adjusted *P* < 0.05 was also applied.

#### Summary-data-based Mendelian randomization (SMR) identifying proteins causally associated with MS

To confirm whether PWAS-identified protein candidates causally influence MS, SMR analysis was performed.^25^ This method tests the pleiotropic association between genetically predicted protein levels and MS onset. A key assumption of this approach is that an identical underlying causal variant determines both protein expression and disease phenotype. However, due to LD, it is possible that the SMR effect could be non-zero even when this assumption is violated. To distinguish pleiotropy from linkage, Heterogeneity in dependent instruments (HEIDI) test was performed, under the assumption that if protein expression and disease phenotype are affected by a shared causal variant, β_SMR_ would be the same for any variant in LD with that causal variant. Thus, greater heterogeneity among β_SMR_ statistics calculated for all cis-pQTLs implies a greater likelihood of linkage, rather than pleiotropy.

SMR and HEIDI analyses were performed using SMR software (version 1.3.1). The 1KG reference panel was used for LD estimation. Common variants with MAF > 0.01 were involved in the analysis. Briefly, for the SMR test, only pQTLs that met the significance threshold of *P*-value < 5×10^−8^ were involved. For HEIDI test, the significance threshold was set to a *P*-value < 1.57×10^−3^; pQTLs were required to correlate with the top-associated cis-pQTL with an *R*^2^ of 0.05-0.90; and the number of involved SNPs was restricted to 3-20. The statistical significance threshold for putative causal proteins was defined using FDR-adjusted *P_SMR_* < 0.05 and *P_HEIDI_* > 0.05.

#### Bayesian co-localization analysis examining shared causal variant influence on both protein expression and MS

To further address the potential effects of LD on the pleiotropic associations, Bayesian co-localization analysis was performed to assess the posterior probability of shared genetic variants being responsible for both protein expression and MS development using the coloc R package (version 5.2.3). In this approach, the posterior probability is generated for the following hypotheses: H0: neither protein expression level nor MS development has a genetic association in the region; H1: only protein expression level has a genetic association in the region; H2: only MS development has a genetic association in the region; H3: both protein expression level and MS development have genetic association but with different causal variants; H4: both protein expression level and MS development have genetic association in the region and share a single causal variant. The posterior probability of H4 (*PPH4*) > 0.75 was considered colocalized.

#### Definition of potential causal proteins

Potential causal proteins were defined as those that successfully passed all three analyses: the SMR analysis (FDR-adjusted *P_SMR_* < 0.05), the HEIDI test (*P_HEIDI_*> 0.05), and the colocalization analysis (*PPH4* > 0.75). These potential causal proteins were recognized as potential drug targets and underwent further comprehensive functional investigations.

#### KEGG pathway annotation, protein-targeted drugs identification, and PPI network analysis

To interpret functional implications, KEGG pathway annotation was performed for all detected potential causal proteins (https://www.genome.jp/kegg/pathway.html). Existing drugs targeting each of the identified potential causal proteins as well as existing MS drug targets were determined by reviewing the Drugbank database (https://go.drugbank.com/). To investigate the interactions between potential drug targets and the known MS drug targets, and to explore the interactions of potential drug targets within and across plasma and brain, PPI network analysis was conducted using the Search Tool for the Retrieval of Interacting Genes database (https://string-db.org/). Only interaction scores greater than 0.4 were shown.

#### SMR identifying mRNA level of detected proteins

To increase the biological interpretability of the identified potential causal proteins, we further examined their mRNA expression levels, investigating compliance with the central dogma of molecular biology. Firstly, for identified potential causal proteins, the corresponding mRNA levels were tested in relation to MS through SMR analysis (integrating eQTL and MS GWAS). Subsequently, for significant MS-associated mRNA expressions, the links between mRNA expressions and protein expressions were investigated through SMR analysis (integrating eQTL and pQTL). The significance threshold was set to *P_SMR_* < 0.05, and probes with *P_HEIDI_* < 0.05 were excluded.

#### Standard Protocol Approvals, Registrations, and Patient Consents

This study was a secondary analysis of existing, publicly available summary-level GWAS and QTL data. The statement of ethics for each research can be found elsewhere, approved by the relevant institutional review board or an equivalent committee^19–23^. Consent to participate: not applicable.

## Results

### Identifying candidate proteins associated with MS through PWAS

To identify candidate proteins linked to MS, we employed PWAS to establish associations between protein abundances in plasma as well as in brain and MS susceptibility. In plasma, we identified 100 proteins whose expression levels were associated with MS susceptibility (*P*-values < 0.05). After FDR correction, 36 out of these 100 proteins remained significant (**Figure 2A, Supplementary Table S1**). Correspondingly, in brain, we identified 212 proteins (*P*-values < 0.05). After FDR correction, 32 out of these 212 proteins remained significant (**Figure 2B, Supplementary Table S2**). The 100 plasma proteins and 212 brain proteins under nominal significance were considered as candidates for subsequent causal inference analysis.

**Figure 2.**
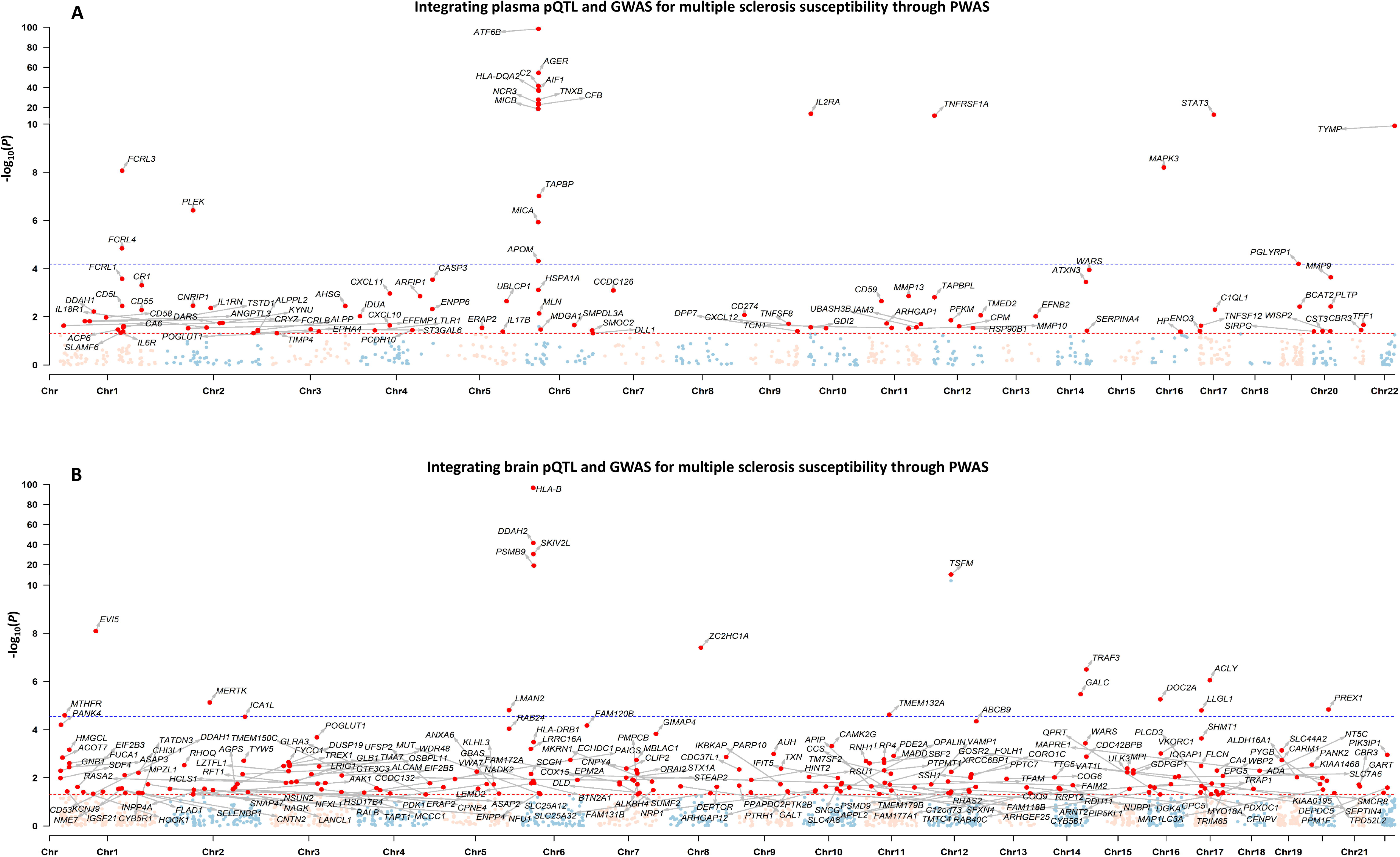
Identifying candidate proteins associated with multiple sclerosis by integrating pQTL and GWAS through PWAS. A: Identifying candidate proteins in plasma associated with multiple sclerosis by integrating pQTL and GWAS through PWAS. B: Identifying candidate proteins in brain associated with multiple sclerosis by integrating pQTL and GWAS through PWAS. The X-axis represents chromosomes. The Y-axis represents the negative logarithm of *P*-values. Each dot on the Manhattan plot represents a gene, whose cis-regulated protein expression level was tested in association with the multiple sclerosis risk. The red dashed line indicates a nominal *P*-value threshold of 0.05, and the blue dashed line indicates the Bonferroni-corrected *P*-value threshold of 6.63×10^−5^ in plasma and 2.86×10^−5^ in brain. Proteins exhibiting nominal significance in association with multiple sclerosis risk were identified as candidate proteins, and their corresponding encoding genes are highlighted with red dots. pQTL: protein quantitative trait loci; GWAS: genome-wide association study; PWAS: proteome-wide association study; Chr: chromosome.

### Identifying proteins causally associated with MS through SMR and colocalization

We used SMR to infer the causal roles of candidate proteins identified by PWAS. In plasma, we identified 57 proteins whose expression levels were causally associated with MS susceptibility (FDR-adjusted *P*-values < 0.05). Among these proteins, 34 exhibited *P*_HEIDI_ > 0.05, indicating that the associations were likely to be pleiotropic rather than impacted by distinct variants in LD (**Figure 3A, Supplementary Table S3**). Using colocalization analysis, we observed 11 proteins with strong posterior probabilities (*PPH4* > 0.75), suggesting a shared causal variant responsible for both plasma protein expression and MS susceptibility, as opposed to distinct causal variants in proximity (**Figure 3A, Supplementary Table S4**). Collectively, we identified nine potential causal proteins in plasma. Among these, highly expressed *CR1 and WARS* were associated with an increased risk of MS, while highly expressed *TNFRSF1A, FCRL3, TYMP, PGLYRP1, CD59, IDUA,* and *ARHGAP1* were associated with a decreased risk of MS (**Figure 3A**).

**Figure 3.**
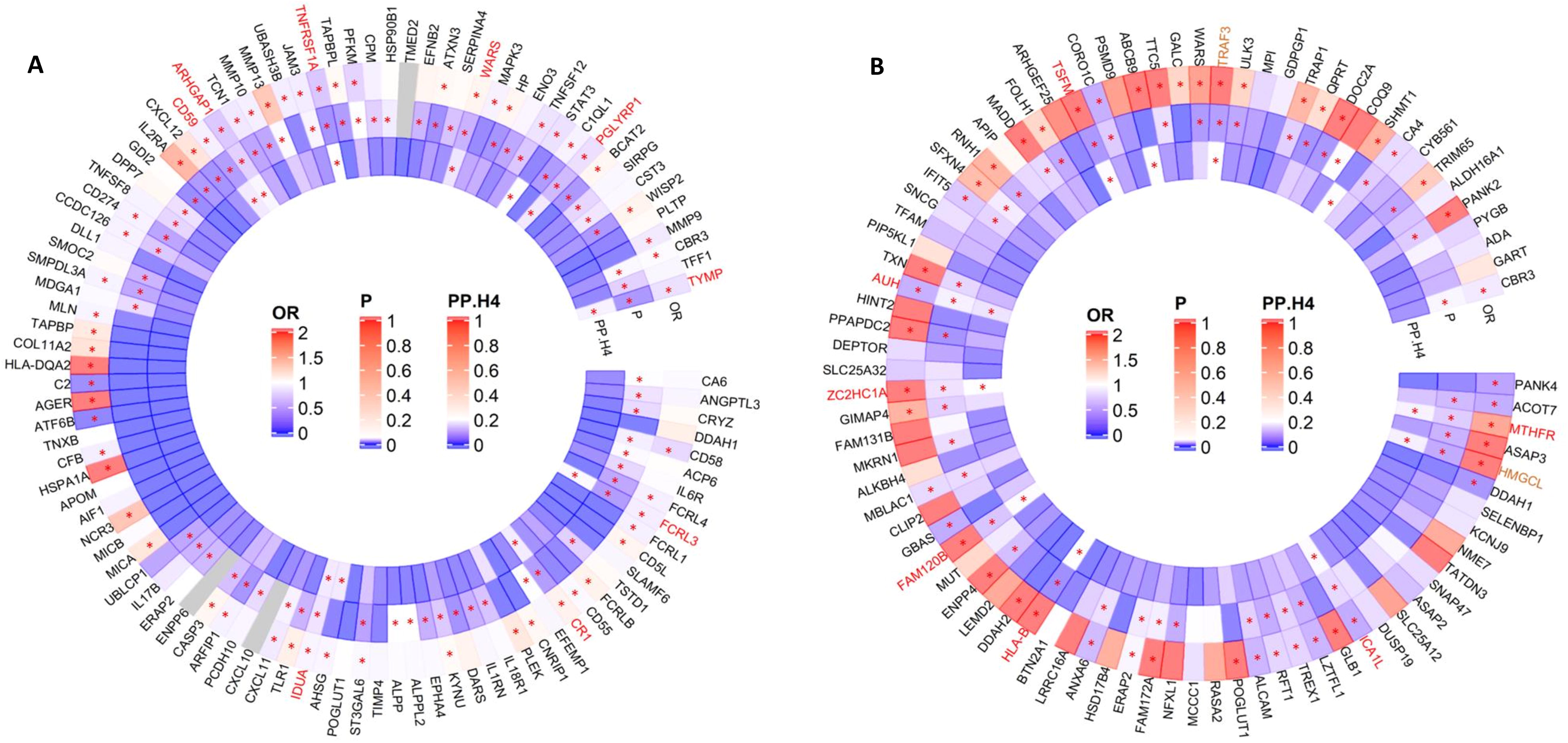
Identifying potential causal proteins for multiple sclerosis through SMR and colocalization analysis, based on candidate proteins determined by PWAS. A: Identifying potential causal proteins in plasma for multiple sclerosis through SMR and colocalization analysis, based on candidate proteins determined by PWAS. B: Identifying potential causal proteins in brain for multiple sclerosis through SMR and colocalization analysis, based on candidate proteins determined by PWAS. The corresponding encoding genes for candidate proteins are listed outside the circle. Odds ratios from SMR analyses are presented in the outer circle. *P*-values for HEIDI tests are displayed in the middle circle. The posterior probabilities for colocalization analysis, under the hypotheses of one shared SNP associated with both protein quantitative traits and multiple sclerosis, are shown in the inner circle. The color of boxes in the circles represents the magnitude of each statistic, with warmer colors indicating stronger effects and gray indicating missing values. Significant results are highlighted with red asterisks, defined as Benjamini-Hochberg False Discovery Rate-adjusted *P* < 0.05 for SMR analysis, *P*_HEIDI_ > 0.05, and posterior probability for colocalization analysis > 0.75. Gene names in red indicate that their corresponding proteins passing all three analyses are identified as potential causal proteins. Gene names in brown indicate that their corresponding proteins are potential causal proteins and adhere to the central dogma. Figure 3B excludes 127 out of 212 PWAS-identified candidate proteins from SMR analysis because their pQTLs had *P*-values > 5×10^−8^, not meeting the SMR instrument variable criterion (pQTLs with *P*-values < 5×10^−8^). The figure illustrates SMR and colocalization results for the remaining 85 out of 212 candidate proteins. SMR: summary-data-based Mendelian randomization; PWAS: proteome-wide association study; OR: odds ratio; HEIDI: heterogeneity in dependent instruments test. PP.H4: posterior probability for colocalization analysis under the hypotheses of one shared SNP associated with both traits.

In brain, we firstly excluded 127 of 212 PWAS-identified candidate proteins from SMR analysis due to their pQTLs having *P-*values > 5×10^−8^, failing to meet the SMR instrument variable criterion (pQTLs with *P-*values < 5×10^−8^). Consequently, from the remaining 85 of 212 PWAS-identified candidate proteins, we identified 48 proteins whose expression levels were causally associated with MS susceptibility (FDR-adjusted *P*-values < 0.05). Among these proteins, 39 successfully passed HEIDI test with *P*_HEIDI_ > 0.05 (**Figure 3B, Supplementary Table S5**), and 11 showed posterior probabilities of *PPH4* > 0.75 via colocalization analysis (**Figure 3B, Supplementary Table S6**). Collectively, we identified nine potential causal proteins in brain. Among these, highly expressed *HLA-B*, *ZC2HC1A*, *HMGCL, TSFM, FAM120B, TRAF3*, and *MTHFR* were associated with an increased risk of MS, while highly expressed *ICA1L* and *AUH* were associated with a decreased risk of MS (**Figure 3B**).

To answer whether associations with MS at protein expression levels also exhibited similar evidence at gene expression levels, we further explored the causal role of mRNA levels of the 18 identified potential causal proteins. For the nine proteins identified in plasma, mRNA levels were profiled in seven. Among these, SMR identified mRNA levels of five genes causally associated with MS susceptibility (*P_SMR_* < 0.05, *P*_HEIDI_ > 0.05, and exhibiting consistent direction of effects as observed at protein levels, **Supplementary Table S7**). Unfortunately, despite these five genes showing evidence at transcriptional and translational levels for MS separately, SMR linking these two molecular traits failed to identify a relationship such that a genetic locus causally affects MS susceptibility through first modifying mRNA expression and then protein translation (**Supplementary Table S8**).

For the nine potential causal proteins identified in brain, mRNA levels were profiled in eight. Among these, SMR identified mRNA levels of three genes (*TRAF3, AUH,* and *HMGCL*) to be causally associated with MS susceptibility (*P_SMR_* < 0.05, *P*_HEIDI_ > 0.05, and exhibiting consistent direction of effects as observed at protein levels, **Supplementary Table S9**). Among these three genes with evidence at transcriptional and translational levels for MS respectively, SMR connecting these two molecular traits suggested that rs2076343 in *HMGCL* and rs7145882 in *TRAF3* were causally associated with MS through modifying mRNA expression and protein translation (**Supplementary Table S10**).

### Pathway annotation, protein-targeted drugs identification, and PPI

The 18 detected potential causal proteins were defined as potential drug targets, none of which are currently known drug targets for MS treatment. To verify our findings, we further performed comprehensive analyses on pathway annotation, protein-targeted drug identification, and PPI to investigate the underlying biological mechanisms, drugability, and interaction network.

For the nine plasma drug targets, 56 pathways were annotated (**Supplementary Table S11**). An intricate interaction network was observed for five of these targets (*CD59, FCRL3, CR1, TNFRSF1A,* and *TYMP*) with the 19 known targets of 11 existing MS drugs including *ublituximab, ofatumumab, ocrelizumab, glatiramer, alemtuzumab, daclizumab, natalizumab, cladribine, interferon beta, dimethyl fumarate,* and *teriflunomide* (**Figure 4, Supplementary Table S12**). Moreover, four of these targets (*IDUA, TNFRSF1A, WARS,* and *TYMP*) were also targeted by 13 existing non-MS drugs, suggesting potential opportunities for drug repurposing (**Figure 4, Supplementary Table S11**).

**Figure 4.**
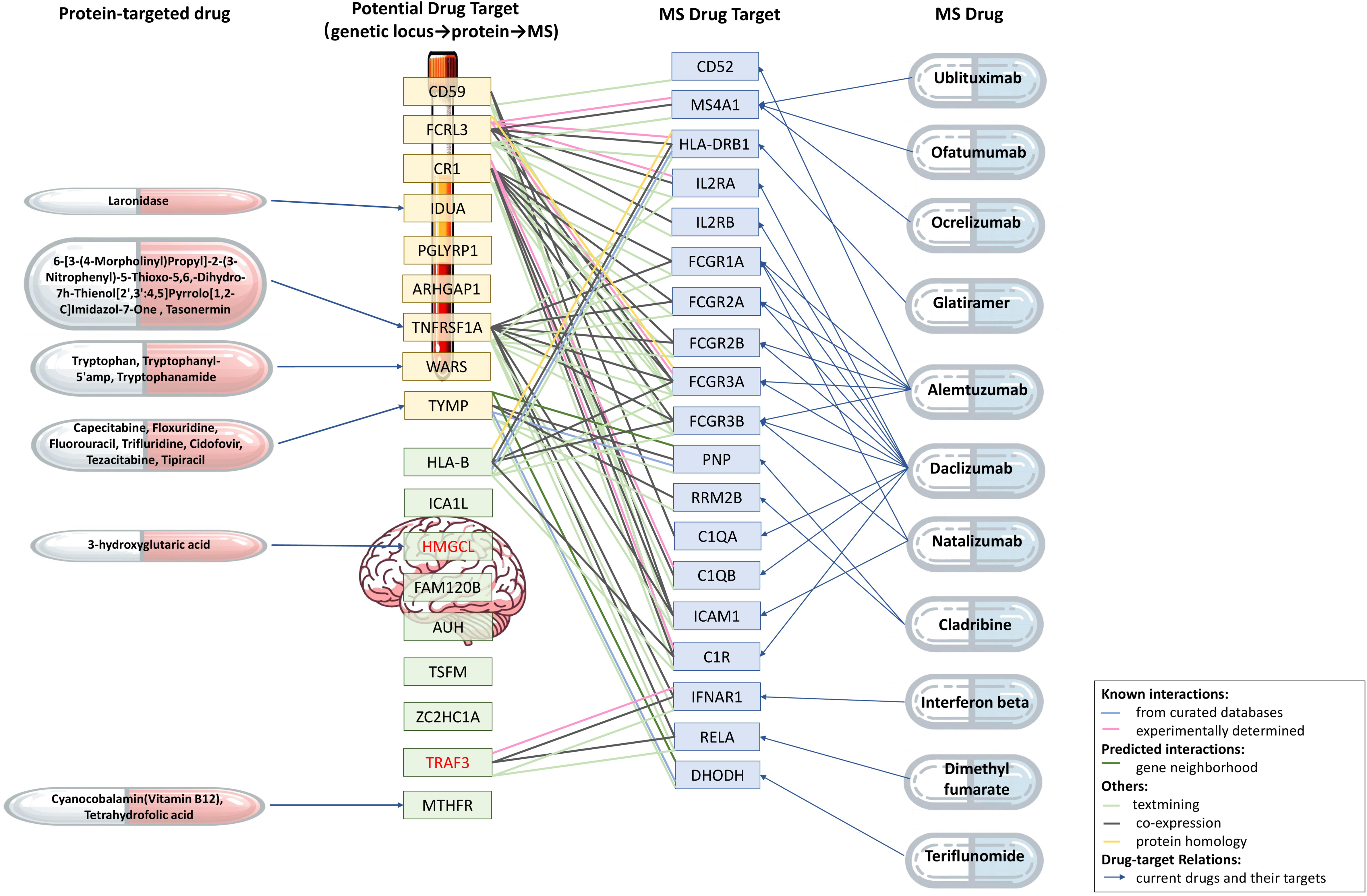
Potential drug targets, protein-targeted drugs, and protein-protein interaction network. The corresponding encoding genes for these potential causal proteins are presented. Gene names in red indicate that their corresponding proteins adhere to the central dogma. Potential drug targets highlighted in yellow boxes are identified in plasma, and those in green boxes are identified in brain. Drug relations refer to the existing drugs targeting the identified potential drug targets. MS drug targets refer to proteins targeted by existing MS drugs. The protein-protein interaction network is presented between potential drug targets and current MS drug targets, with lines of different colors representing various types of interaction. MS: multiple sclerosis; HEIDI: heterogeneity in dependent instruments test.

For the nine brain drug targets, 36 pathways were annotated (**Supplementary Table S13**). An intricate interaction network was observed for two of these targets (*HLA-B* and *TRAF3*) with six known drug targets of five existing MS drugs, including *glatiramer*, *daclizumab*, *alemtuzumab*, *interferon beta*, and *dimethyl fumarate* (**Figure 4, Supplementary Table S14**). Moreover, two of these targets (*HMGCL* and *MTHFR*) were also targeted by three existing non-MS drugs, providing a source of drug repurposing (**Figure 4, Supplementary Table S13**).

### PPI analysis among identified potential drug targets in plasma and brain

A PPI network analysis was conducted encompassing all 18 detected potential drug targets (**Figure 5, Supplementary Table S15**). Among these, six in plasma and five in brain met a minimum interaction score of 0.4. Comprehensive interactions were observed within plasma and brain. Particularly noteworthy were the interactions observed across plasma and brain (*TNFRSF1A-TRAF3* and *WARS-TSFM*).

**Figure 5.**
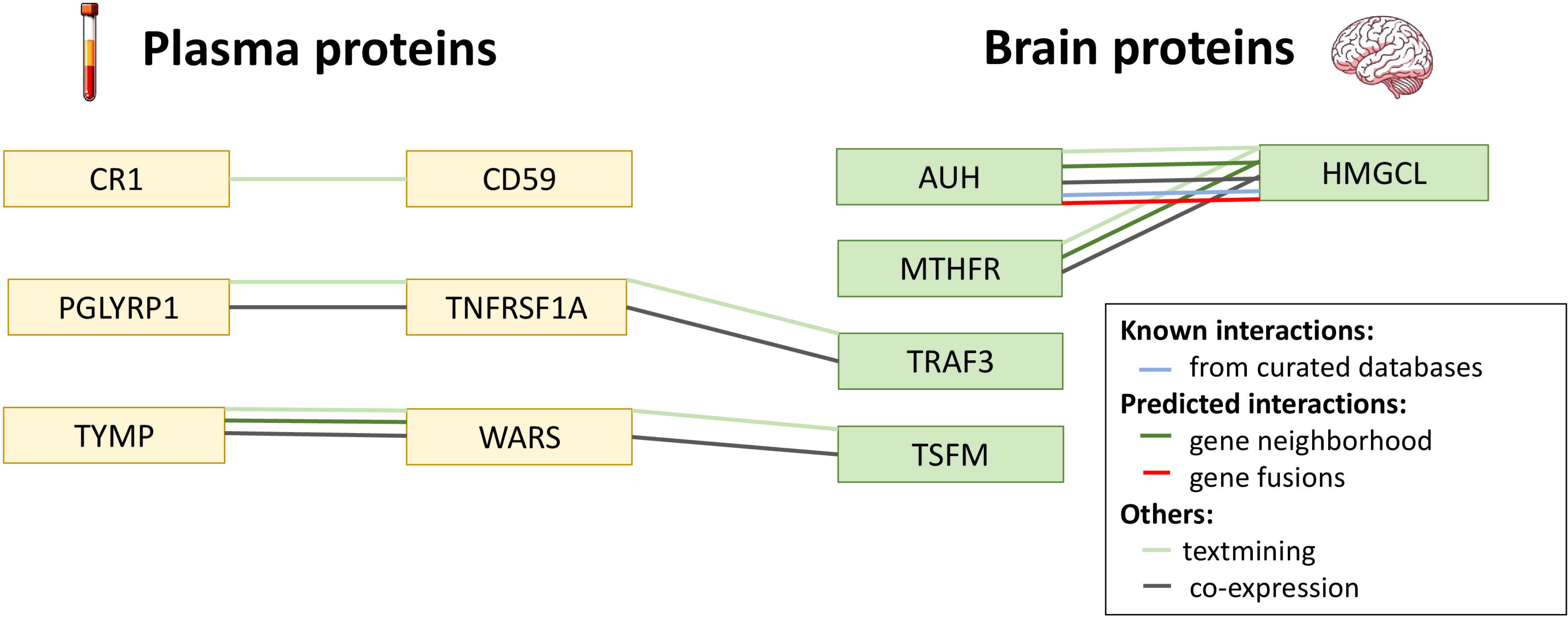
Protein-protein interaction network analysis among all identified potential drug targets in plasma and brain. Protein-protein interaction network analysis was conducted among all detected potential drug targets (nine in plasma and nine in brain). Only those with a minimum required interaction score of 0.4 (six in plasma and five in brain) are considered interaction and presented in this figure. The corresponding encoding genes for these potential causal proteins are presented. Potential drug targets in yellow boxes are identified in plasma, and those in green boxes are identified in brain. The different colored lines connecting these potential drug targets represent various types of interactions.

### Identifying additional druggable proteins through relaxed criteria

As both HEIDI and colocalization analysis aimed to determine a shared causal variant responsible for multiple traits, to identify an enlarged number of potentially druggable proteins for MS, we relaxed the criteria, requiring passing SMR together with either HEIDI or colocalization test, rather than both HEIDI and colocalization test. A total of 78 proteins (37 in plasma and 41 in brain) successfully passed the relaxed criteria (**Supplementary Table S16-17**). Among these, two proteins (*CBR3*, *WARS*) were identified in both plasma and brain. Half (39 of 78) identified proteins were confirmed by previous epidemiology or laboratory studies, proving the reliability of these findings. Moreover, 13 of 78 (one in plasma and 12 in brain) proteins adhere to central dogma (**Supplementary Table S18-S21**).

For the 37 proteins identified in plasma, 22 were annotated in 127 biological pathways. In total, 17 proteins were targeted on 62 existing non-MS drugs, and 18 proteins showed interaction with 23 known targets of existing MS drugs. Importantly, one of the identified proteins *IL2RA* was a known MS drug target (**Supplementary Table S16 and S22**). For the 41 proteins identified in brain, 20 were annotated in 69 biological pathways. In total, 10 proteins were targeted on 45 existing non-MS drugs, and 11 proteins showed interaction with 15 known targets of existing MS drugs (**Supplementary Table S17 and S23**). Intricate PPIs among the 78 proteins were identified (**Supplementary Table S24**).

For the 13 potential drug targets adhering to the central dogma, six were annotated in 30 biological pathways. In total, two proteins were targeted on four existing non-MS drugs, and three proteins interacted with three known targets of existing MS drugs (**Supplementary Table S16-S17, and Supplementary Figure S1**).

## Discussions

In our study, we tested hundreds of MS-associated proteins in plasma and brain and confirmed the potential causal role of 18 proteins through comprehensive analytical strategies. None of these are currently known MS drug targets, we therefore defined these as potential novel drug targets. We revealed intricate interactions between seven of these 18 proteins and 19 known MS drug targets, as well as interactions among 11 of these 18 proteins within and across plasma and brain. Furthermore, we identified 16 existing non-MS drugs targeting six of these 18 potential targets. These findings present significant potential for both the discovery of new drugs and the repurposing of existing ones. The reliability of our findings was further supported by 78 annotated pathways and transcriptional evidence on two targets. In the contemporary pharmaceutical industry, the development of new drugs is time-consuming, costly, and highly vulnerable to termination. On average, it takes about 13 years, with costs from hundreds of thousands to over a billion US dollars and a failure rate exceeding 90% before reaching the market.^26^ Our study integrating large-scale omics data using cutting-edge statistical approaches helps prioritize drug targets by elucidating pathological mechanisms, thus may improve the effectiveness of new drug development.^11^

Proteins, as the most common and highly effective drug targets, have been investigated in MS field in two previous integrative studies. Our study largely replicated these findings, successfully confirming six out of nine plasma proteins identified by the previous studies under the relaxed criteria ( *CD40* was no corresponding pQTL in our panel). Moreover, *FCRL3, TYMP, CR1, CD59,* and *TNFRSF1A* were replicated even under the stringent criteria. By incorporating the hitherto largest number of proteins (2,004 in plasma and 1,443 in brain) and utilizing PWAS in combination with SMR to reduce statistical burden, our study extends previous work by identifying the largest numbers of MS drug targets to date (18 proteins passed stringent criteria and 78 proteins passed relaxed criteria). This significantly advances our understanding of MS pathology and enhances the potential for future drug development. Considering the pathogenesis of MS in the central nervous system, our study also identified proteins in brain tissue. These potential causal brain proteins advance the associative findings of the previous study,^18^ thereby expanding the treatment perspective directly to the most relevant tissue. Another advantage of our research is the involvement of transcriptional evidence for the identified potential drug targets, enhancing understating of the MS molecular mechanisms. Notably, two potential protein drug targets (*HMGCL* and *TRAF3*) adhered to the central dogma, greatly strengthening their credibility for further preclinical and clinical development.

We prioritize seven potential drug targets with identified biological relevance connected to known MS drug targets. As *FCRL3*, *TNFRSF1A*, *CR1, CD59,* and *TYMP* have already been discovered and confirmed by the two previous integrative studies,^14,15^ we therefore emphasize two novel findings. ***HLA-B*** is a major histocompatibility complex (MHC) class I molecule presenting antigens to T cells. In alignment with our findings, protective effects of *HLA-B* against MS susceptibility have been observed in *HLA-B*44:02*, *HLA-B*38:01*, and HLA-B*55:01.^27^ *HLA-B* has been annotated to many pathways, such as natural killer cell mediated cytotoxicity,^28^ human cytomegalovirus infection,^29^ and Epstein-Barr virus infection,^30^ all of which have been implicated in MS development. Our study identified strong interactions between HLA-B and HLA-DRB1, a known target of *Glatiramer*, suggesting that both proteins are involved in the same biological processes relevant to MS and reinforcing *HLA-B* as a potential target. ***TRAF3*** regulates NF-kappa-B pathway^31^ which mediates inflammation in MS.^32^ Its other annotated pathways, such as NOD-like receptor signaling, RIG-I-like receptor signaling, IL-17 signaling, and TNF signaling, significantly impact autoimmune disease and MS development.^33–35^ Previous studies have presented that *TRAF3* degradation promotes microglia-mediated CNS inflammation,^36^ and *TRAF3* haploinsufficiency syndrome exhibits B cell hyperactivity leading to hypergammaglobulinemia and autoimmunity.^37^

The 11 potential drug targets that do not interact with known MS drug targets reveal novel biology, presenting opportunities for the development of new drugs. Here, we highlight two potential drug targets with strong supporting evidence. ***IDUA*** is responsible for the glycosaminoglycans heparan sulfate and dermatan sulfate degradation. Its annotated pathways of glycosaminoglycan degradation,^38^ metabolic pathways,^39^ and lysosome function^40^ have been proven to play a pivotal role in MS development, underscoring a promising potential target. ***PGLYRP1*** is an innate immunity protein with functions in antimicrobial and antitumor defense systems. Although no pathway is currently annotated to *PGLYRP1*, its potential in immunotherapy has been proven by a study demonstrating *PGLYRP1* as a proinflammatory molecule in myeloid cells during autoimmune conditions.^41^ Additionally, elevated levels of *PGLYRP1* protein have been observed in the white and gray matter of cerebellum and spinal cord in patients with MS.^42^

Drug repurposing accelerates the translation of scientific discoveries into clinically beneficial treatments. Among the 16 identified non-MS drugs, we first would like to highlight eight existing non-MS drugs including *Tasonermin*, *Tipiracil*, *Capecitabine*, *Floxuridine*, *Fluorouracil*, *Tezacitabine*, *Trifluridine*, and *Cidofovir*, the targets of which interact with known MS drug targets, suggesting the possibility for these non-MS drugs to benefit or assist the treatment of MS. Among these non-MS drugs, the first six possess cancer-therapeutic properties and the last two are antiviral agents. However, these non-MS drugs need prescriptions, and are usually with higher toxicity or stronger side effects compared to over-the-counter drugs, making repurposing difficult. We next would like to highlight vitamin B12 and tetrahydrofolic acid (a folic acid derivative), both are easily obtainable and cost-effective drugs, targeting the protein *MTHFR* (a potential MS drug target). Studies have demonstrated that both vitamin B12 and folate improve myelin regeneration and have neuroprotective and immunomodulatory properties.^43, 44^ Moreover, supplementing vitamin B12 and folate may help reduce homocysteine levels,^45, 46^ a neuro- and vascular-toxic sulfur-containing intermediary product that has been proven to be elevated in MS patients.^47^ Although some studies have provided inconsistency findings,^48, 49^ a clinical study has demonstrated that supplementation with folic acid and vitamin B12 improves the quality of life in MS patients.^45^ Considering the outcome of our study on MS susceptibility, supplementation with vitamin B12 and folic acid may serve as prophylactic agents against the onset of MS. Additionally, combining vitamin B12 and folic acid with DMTs may offer an effective approach for MS management. These potential repurposed drugs could accelerate the clinical process by reducing drug developmental time and costs as well as by leveraging known safety and side effect profiles.

The interactions of potential drug targets across plasma and brain suggest the possibility of substance transport or signaling molecules across blood and brain, which could indicate that targets identified in plasma may influence MS pathology in the brain. This has been validated by results based on relaxed criteria, through which we identified two potential proteins (*CBR3* and *WARS*) present in both plasma and brain tissue, suggesting the possibility of a link between systemic circulation and neurological processes. Furthermore, the limited overlap between targets identified in plasma and brain could be attributed to their distinct protein compositions, cellular functions, and the restrictive nature of the blood-brain barrier.

The interpretation of our study should be approached with caution. First, not every protein has pQTLs. For example, in our referenced plasma pQTL study, less than half of the proteins (45%, 2,004 out of 4,435) possessed significant cis-pQTLs.^19^ Consequently, using pQTLs in integrative analysis, rather than relying on quantitative proteomic analysis, results in the omission of proteins. Nevertheless, integrative analysis allows for making causal inferences, representing an improvement over observational association design. Furthermore, our study has identified the hitherto largest number of potential MS drug targets owing to the large sample size of pQTL data, representing an advantage over the small sample sizes that usually characterize the current quantitative proteomic analysis. Second, among the 18 potential drug targets, only two showed evidence at both transcriptional and translational levels. The absence of transcriptional level evidence for most drug targets could be due to post-transcriptional regulation or protein function through post-translational modification, which, from another aspect, highlights the importance and superiority of a direct investigation on the proteome. Nevertheless, evaluating the expression of individual proteins does not fully reflect the complexity of cellular processes, thus our potential drug targets identified in silico are not conclusive. Further evidence in vitro and in vivo is necessary to validate these findings.

## Data Availability statement

All the genetic and QTL data used in this study were from the publicly accessible summary statistics and can be accessed through the corresponding references presented in the main text.^19–23^

## Acknowledgements

We would like to express our appreciate to all investigators and consortium for sharing the summary statistics.

## Declaration of interests

IK has received lecture honoraria from Merck. None concerns the present paper.

TO has received lecture/advisory board honoraria and unrestricted MS research grants from Biogen, Merck, and Novartis. None of which concerns the present paper.

LA has received lecture honoraria from Biogen and Merck. None of which concerns the present paper.

## Contributors

YJ and QL analysed the data and interpreted the results. YJ wrote the manuscript. YJ and XJ designed the study. QL, PS, IK, TO, LD-G, and LA provided statistical expertise and support in their disciplinary field. XJ, QL, PS, IK, TO, LD-G and LA revised the manuscript. All authors provided the corresponding author with permission to be named in the manuscript. XJ is the guarantor of this study. The authors read and approved the final version of the manuscript.

## Funding

IK were funded by Horizon Europe WISDOM project (grant no 101137154). IK has received academic grants from The Swedish Research Council and the Swedish Brain Foundation.

TO has received academic grants from The Swedish Research Council, the Swedish Brain Foundation, Knut and Alice Wallenberg Foundation, and Margaretha af Ugglas Foundation.

LA has received academic grants from The Swedish Research Council, The Swedish Council for Health, Working Life and Welfare, the Swedish Brain Foundation, and Region Stockholm.

XJ is supported by a starting grant of Karolinska Institutet (2-1534/2020).

Open Access funding was provided by Karolinska Institutet.

## Notes

### Author Declarations

The study used (or will use) ONLY openly available human data that were originally located at:http://nilanjanchatterjeelab.org/pwas/ https://www.synapse.org/Synapse:syn23627957/ https://imsgc.net/ https://yanglab.westlake.edu.cn/software/smr/#DataResource

